# Cost-Effectiveness of an Integrated Antenatal Care Testing Panel to Accelerate the Triple Elimination of Mother-to-Child Transmission of HIV, Syphilis, and Hepatitis B in Nigeria: Modeling Study

**DOI:** 10.64898/2026.08.14.26360312

**Authors:** Isaiah K. Dzimbiri, Pacifique Dusabeyezu, Abdalrahman Ahmed, Missiani Ochwoto, Michael Kingsley, Donald S. Shepard

## Abstract

**Background:** In 2025, the World Health Organization pre-qualified an integrated antenatal care (ANC) testing panel that tests for HIV (including p24 antigen and antibody), syphilis, and hepatitis B (HBV) with one finger prick. The panel would accelerate the triple elimination of vertically transmitted infections by shortening the HIV detection window and increasing testing rates. Nigeria is considering adoption but needs performance and cost projections.

**Methods:** We constructed deterministic (with Microsoft Excel) and probabilistic (with Python) models, including parallel testing and treatment post-exposure prophylaxis algorithms for positive p24. We calibrated the models to Nigeria’s 6.4 million women entering ANC annually using epidemiologic literature, product prices, and occasionally expert opinion. We compared costs (in 2025 US dollars) and outcomes between current and projected future (2027) practices.

**Results:** The panel would avert 562 of the current 3,299 vertical infections per 100,000 women in ANC. Per woman in ANC, the panel would avert 0.0656 disability adjusted life years (DALYs) at a net cost of US$7.55. With low current testing rates. HBV testing averts the most DALYs (33%), followed by acute HIV (29%), chronic HIV (25%), and syphilis (14%). The incremental cost-effectiveness ratio (ICER) is $115 (95% confidence interval: $91-$143) per DALY averted--more favorable than Nigeria’s conservative historical $137 average. The benefit-cost ratio is also favorable (1.19; 95% confidence interval: 0.93-1.51).

**Conclusions:** The integrated ANC testing panel would be a valuable and cost-effective addition to ANC care. Piloting in Nigeria and similar sub-Saharan African countries would refine parameters for potential scale up.

## BACKGROUND

### Triple elimination

In 2023, the World Health Organization published a framework for implementing triple elimination of mother-to-child transmission (EMTCT) of HIV, syphilis and hepatitis B (HBV)[1]. Its key elements are testing in antenatal care (ANC) clinics and prompt and efficacious interventions to treat women who test positive.

In response to low testing rates for syphilis and HBV, WHO prequalified an integrated ANC testing panel in 2025. This panel integrates detection of p24 antigen, HIV antibody, syphilis, and HBV into a single package[2]. The rapid tests use lateral-flow assays from a single finger-stick blood sample. This can reduce workload, extend existing HIV program infrastructure to syphilis and HBV, and lower out-of-pocket costs and time burden for pregnant women. Additionally, the panel’s inclusion of p24 antigen, complementing antibody-only tests in previous products, allows HIV infections to be detected 14 days earlier in pregnancy. Because the panel targets a phase of very high viral load and 26-fold elevated risk of HIV vertical transmission[3], it offers an important advantage in helping to stop HIV transmission[1,4]. This approach extends the commitment, approaches and public financing previously dedicated to HIV services to all three infections and streamlines inventory management. The panel was announced at the International AIDS Society 2025 conference in Kigali, Rwanda[5].

Nigeria, the most populated country in Africa, bears one of the largest global burdens of HIV, syphilis and chronic HIV among pregnant women[6]. While Nigeria has achieved relatively high levels of testing for HIV, screening rates for syphilis and HBV testing remain comparatively low, leaving many pregnant women and their infants at risk of the adverse effects of preventable infections. Another challenge is the current testing for HIV is based on antibodies. Nigeria is considering incorporating this integrated ANC testing panel into its ANC program. With the termination of USAID in 2025 and scaling down of other donors in Nigeria and across sub-Saharan Africa, costs are a major concern. Yet, to our knowledge, no economic studies of the integrated ANC testing panel exist.

### Study goals and objectives

To address this gap, we developed and performed an economic appraisal of the integrated ANC testing panel. Our first objective was to develop and apply deterministic and probabilistic decision-analytic models. They assess the cost-effectiveness of introducing the WHO-prequalified integrated ANC testing panel for HIV, syphilis and HBV in Nigeria for the projected 2027 ANC cohort, compared with continuation of current testing practices. The model estimates incremental infections detected and disability-adjusted life years (DALYs) averted, programmatic and treatment costs, and the incremental cost-effectiveness ratio (ICER) from a health-care sector perspective. Based on literature, our base model treats p24-reactive results analogous to post-exposure prophylaxis (PEP) for acute HIV infection. Our second objective was to conduct a benefit-cost analysis of the testing panel. Our final objective was to compare our base model against alternative algorithms for positive p24.

## METHODS

### Model structure

Diagnostic tests do not directly prevent or treat infections; instead, they provide information that enables health systems to enhance prevention and treatment strategies applying a specified algorithm. Assessment also requires projections of real-world behaviors of providers and patients, based on their knowledge, costs, and the availability of products and services.

We created both deterministic and probabilistic versions of our model. The deterministic version was structured as an economic and epidemiologic workbook, containing 60 interrelated Microsoft Excel sheets (supplement 1). Of these, 47 contained data and formulas, while the remaining 13 are purely informational (references and descriptive material). The Excel model builds on one previously developed including malaria for three East African countries[7]. The probabilistic version, designed to perform sensitivity analyses, recreated the mathematical relations from the Excel workbook in Python (supplement 2). Using the range (from the literature) and the chosen distributions for each parameter (normal for continuous and beta for probability values) the Python version was used to perform 10,000 iterations. These generated probability distributions on the key results.

### Infection prevalence rates

We determined the four prevalence rates (acute HIV, chronic HIV, syphilis, and HBV) from the literature. Our best estimate was the average of relevant studies. Our sensitivity analysis was based on the confidence interval or range of the underlying sources. We used data from Nigeria where possible. However, for acute HIV, where we found no relevant Nigerian study, we incorporated relevant literature from sub-Saharan Africa more widely using a Kenyan study [8]. For chronic HIV, we relied on the mean and standard deviation for a key subset of a 2018 national sample[9]: the 7,039 females aged 15-49 who were pregnant at the time of the study. For syphilis, where we had multiple original studies, we took the second from the lowest and from the highest values to define our range. For HBV, where our analysis was based on three regions (northern, southern, and eastern), our best estimate was the population-weighted mean of these regions. Our extremes were based on the lowest and highest regions. For HBV, the transmission risk depends on the birth dose vaccination, treatment of the infected infant with immunoglobulin, and short-term treatment (through 24 weeks postpartum) of the pregnant woman with tenofovir. Future practice has higher, but not complete, probabilities of the recommended steps. Supplement 3 gives the central and alternative values and sources for prevalence and all other model parameters.

### Screening and treatment rates

Current screening and treatment rates were based on internal program data. Currently, Nigeria combines single tests for each infection and a dual test that detects both HIV and syphilis in a single rapid test. Future screening is based on the introduction of the ANC testing panel with rollout based on feasibility, costs, and health benefits. The ANC testing panel was projected to replace the dual HIV and syphilis test. However, the single HIV antibody test was projected ton continue due to existing stocks and procurement arrangements.

Treatment was projected first on pregnant women identified as positive (reactive) after initial and, as appropriate, confirmatory tests. Treatment rates were then computed on a cohort basis as the share of infected members of the cohort being treated, combining screening and treatment of identified persons. Net impacts were the differences between future and current practices.

### Costs of tests and treatment

Unit costs of test products came from published prices or procurement records. Our cost analysis wis conducted in constant 2025 US dollars. Thus, the same unit costs were applied for both current and future conditions. Staff time (technician, nurse, receptionist, and a supervisor) was based on a time simulation of testing with actual staff, plus a staff member simulating the role of a pregnant woman in ANC, at Mararaba Primary Healthcare Center, Nasarawa State, in May 2025 led by one of the authors (MK).

As the integrated ANC testing panel is a single product with an overall acquisition price, we separated testing costs into registration and implementation components. The registration component was the price of the integrated panel. The implementation cost for each component summed confirmation testing, as needed, offering treatment based on test outcomes and guidelines, and treatment receipt based on projected practices, and outcomes based on the literature.

### Probabilistic sensitivity analysis

We performed a probabilistic sensitivity analysis incorporating the variability in prevalence rates and net cohort-based treatment rates. For prevalence rates, where we had specific central, lower and upper uncertainty intervals, we used PERT (Performance evaluation review technique) to create the distribution. For net cohort treatment rates, we estimated moderate lower and upper bounds that were 25% below and above, respectively, the central value. We selected this moderate variability because the short time to future conditions made major health systems changes unlikely. We considered each parameter to be statistically independent of others.

We used Claude (an AI coder) to help create, review, and the Python version of the model. We validated the resulting distribution of ICERs and benefit-cost ratios by confirming that the medians from Python agreed with our original Excel deterministic analysis. We displayed the resulting sensitivity analysis through histograms in the ICER, the benefit-cost ratios, and box plots on the ICER for each infection and overall.

### Alternative algorithms for positive p24

As testing for p24 antigen is not yet in routine use in ANC in Nigeria, no testing algorithm has been publicly released. If a woman tested positive for both antigen and antibody (which happened in the Kenya study due to overlapping reference periods) existing antibody protocols were applied. Alternative protocols were considered for pregnant women in ANC who tested negative for antibody but positive for antigen (p24). Based on preliminary estimates, our basic option (Provisional ART) entailed parallel treatment and testing, leveraging the algorithm for post-exposure prophylaxis. Provisional ART entails initiating a provisional dose of ART at the same visit in which the woman first tested positive for p24. Under this option, she is requested to return in 4 weeks for follow-up testing and with her treatment updated based on the follow-up results. The next alternative (4-week return) provides no provisional ART and asks her to return in 4 weeks for repeat testing. The next alternative (StatPak, 4-week return) uses the StatPak for confirmation and asks the woman to return in 4 weeks for re-testing if negative. The next alternative (StatPak, PCR) conducts polymerase chain reaction (PCR) if StatPak is negative. The final alternative (PCR referral) refers the woman for PCR testing after a positive p24. Each option strikes different balances between the potential benefits of early treatment against costs, feasibility, and risks. The model recommends the alternative with greatest net economic benefit[10].

### Ethical approval

The study was approved by the Nasarawa State (Nigeria) research ethics committee in May 2025. No identified data were accessed or used.

## RESULTS

### Screening rates

The number of women screened under current and future conditions by infection is shown in Figure 1. Currently, no screening test for acute HIV (based on p24 antigen) is used in ANC screening in Nigeria. In future practices, we project that the ANC testing panel will be used for 67% of women in ANC as the ANC panel was projected to replace the dual HIV/syphilis test. For acute HIV, the number of women screened thus increased from 0 (under current conditions) to 4.29 million women (under future conditions), the most dramatic change.

**Figure 1:**
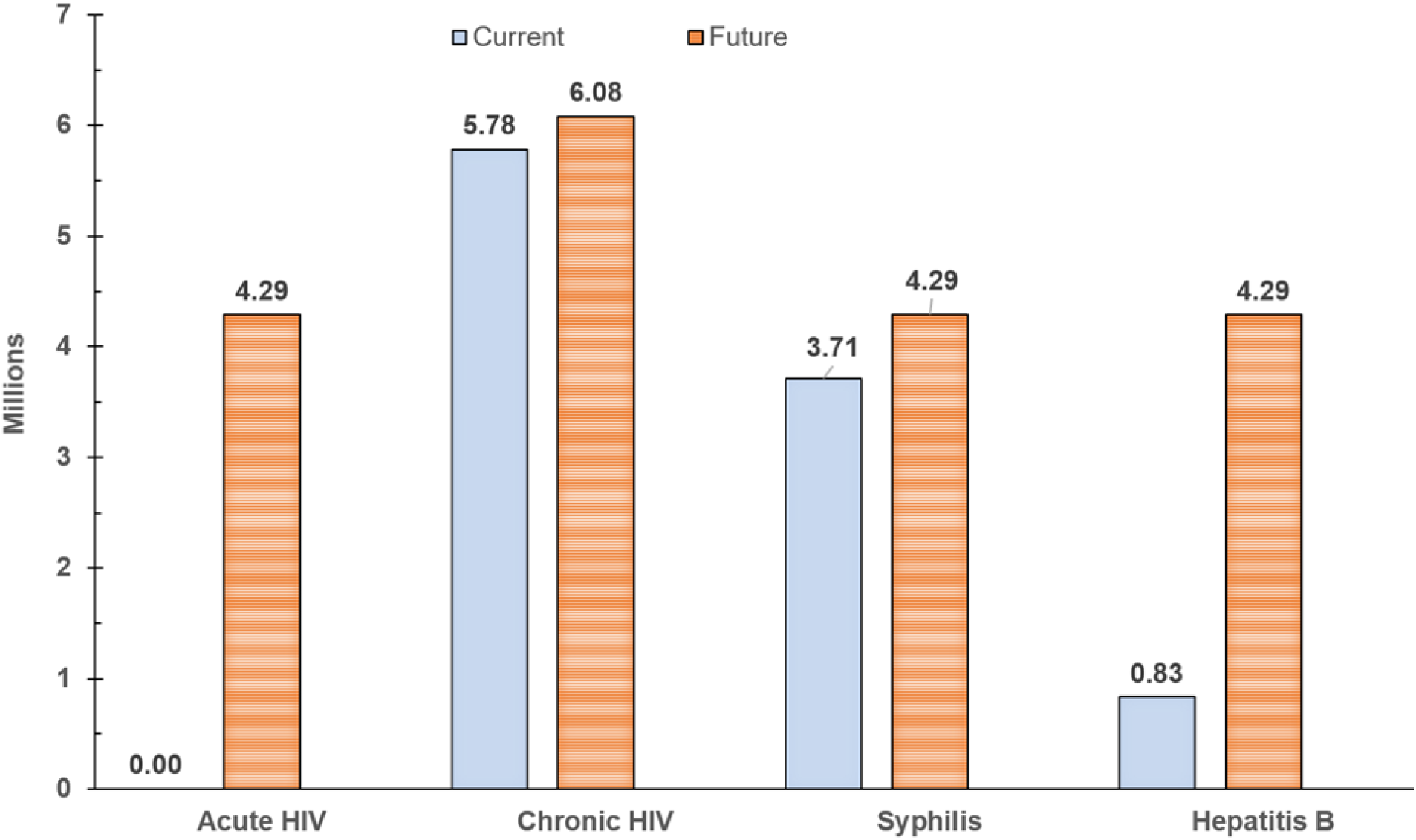
Number of women screened from 6.4 million entering antenatal care annually

Chronic HIV, which had the highest current screening rate (90.4%) of any infection (5.78 million women in ANC), was projected to increase to 95%, (6.08 million women). While the numerical increase (0.30 million) is relatively small, chronic HIV would remain the most widely tested infection and would reach a very high share of the eligible cohort. Syphilis testing would see a moderately large increase as the ANC testing panel is projected to replace the dual test. HBV screening, being part of the ANC testing panel, would also see a major increase.

### Other model inputs

Reflecting uncertainty around rates of disease, screening and treatment, The upper panel Table 1 provides best estimates, lowest, and highest values for key epidemiological parameters. For p24, we found only one usable peer reviewed publication reporting prevalence in ANC [8]. As only 3 of the 577 pregnant women in the study were positive for p24 but negative for HIV antigen, the estimated prevalence is 0.070%. However, the range of uncertainty was substantial. The highest estimate (0.28%) was 16.5 times the lowest (0.017%) for acute HIV and 1.6 times the lowest for chronic HIV (1.40% vs. 0.910%, respectively). HBV is the most prevalent infection (3.84%). As prevalence rates reflect underlying disease characteristics, they were considered the same for both current and future conditions.

**Table 1:** Key Model Inputs by Infection*.

| Row | Input | HIV<br>(p24) | Chronic<br>HIV | Syphilis | Hepatitis<br>B |
| --- | --- | --- | --- | --- | --- |
| <u>Rates of disease, screening, and treatment</u> |  |  |  |  |  |
| (1) | Prevalence (best estimate) | 0.070% | 1.100% | 1.570% | 3.840% |
| (2) | Prevalence (lowest estimate) | 0.017% | 0.910% | 0.380% | 1.600% |
| (3) | Prevalence (highest estimate) | 0.280% | 1.400% | 2.710% | 5.610% |
| (4) | Screening rate, current | 0.0% | 90.3% | 58.0% | 13.0% |
| (5) | Dual (HIV & syphilis) | 0.0% | 52.0% | 52.0% | 0.0% |
| (6) | Single | 0.0% | 38.3% | 6.0% | 13.0% |
| (7) | Screening rate, future | 67.0% | 95.0% | 73.0% | 67.0% |
| (8) | Antenatal triple panel | 67.0% | 67.0% | 67.0% | 67.0% |
| (9) | Dual | 0.0% | 0.0% | 0.0% | 0.0% |
| (10) | Single | 0.0% | 28.0% | 6.0% | 0.0% |
| (11) | Tx rate (identified), current | 0.0% | 92.0% | 55.0% | 7.5% |
| (12) | Tx rate (identified), future | 95.0% | 95.0% | 60.0% | 30.0% |
| (13) | Tx rate (cohort), current | 0.0% | 83.1% | 31.9% | 1.0% |
| (14) | Tx rate (cohort), future | 63.7% | 90.3% | 43.8% | 20.1% |
| (15) | Tx rate increase (cohort) | 63.7% | 7.2% | 11.9% | 19.1% |
| <u>Unit costs</u> |  |  |  |  |  |
| (16) | Single tests: current cost by component | \$0.00 | \$0.90 | \$0.90 | \$0.90 |
| (17) | Dual HIV/syphilis test: Allocated cost by component | \$0.00 | \$0.475 | \$0.475 | n.a. |
| (18) | Confirmatory HIV test (Uni-gold) | \$1.60 | \$1.60 | \$0.00 | \$0.00 |
| (19) | Lifetime treatment per infant | \$31.89 | \$31.89 | \$10.00 | \$30.00 |
| (20) | Lifetime treatment per mother (20.77 discounted years) | \$2,783 | \$2,783 | \$10 | \$154 |

The lower panel of Table 1 shows unit costs of tests and treatment. As noted, while screening helps to identify women with an infection who may benefit from treatment, only treatment of the and/or their infants can generate health benefits. However, we project that broadening screening for HBV and syphilis will sensitize both pregnant women and health workers around the need for treating those infections, thereby increasing the treatment rate among those identified. To reflect this synergistic benefit, we also show treatment rates on a cohort basis (the share of those infected being treated), calculated as the product of the screening and identified treatment rates. For our cost-effectiveness analysis, the percentage point increase from current to future conditions is more is generally the major driver of costs and outcomes. This percentage point increase in the cohort treatment rate was greatest for acute HIV (63.7%) followed by HBV (19.1%).

As noted, the model has only a single value for both current and future unit costs when expressed in constant US dollars, as these come from public sources with negligible uncertainty. The health contributions of the ANC testing panel towards each infection are the combined effect of its technical performance on detection and its health system performance for pregnant women and health workers in accessing and maintaining treatment.

### Health benefits

Table 2 shows projected vertical transmissions and DALYs averted. The ANC testing panel and associated treatment will avert 562 more vertical transmissions per 100,000 women in ANC than the 820 under current testing and treatment, a 69% increase. The distribution shows that these are primarily due to prevention of HBV (76%) and syphilis (16%). These results are consistent with the substantial increase in screening for these two infections. The health benefits (DALYs averted) of detection and treatment for HBV are 33% of the total DALYs averted in the cohort. Across all infections, the ANC testing panel and associated treatment avert 419,577 DALYs. Dividing this benefit by the cohort size (6,400,000 women) gives a substantial benefit of 0.0656 DALYs averted. This number shows that improving detection of infections in pregnancy would be a powerful public health intervention. The DALYs averted, which benefit both the woman (63%) and the infant (37%), are equivalent to about one month of illness or premature death avoided, per woman in ANC.

**Table 2:**
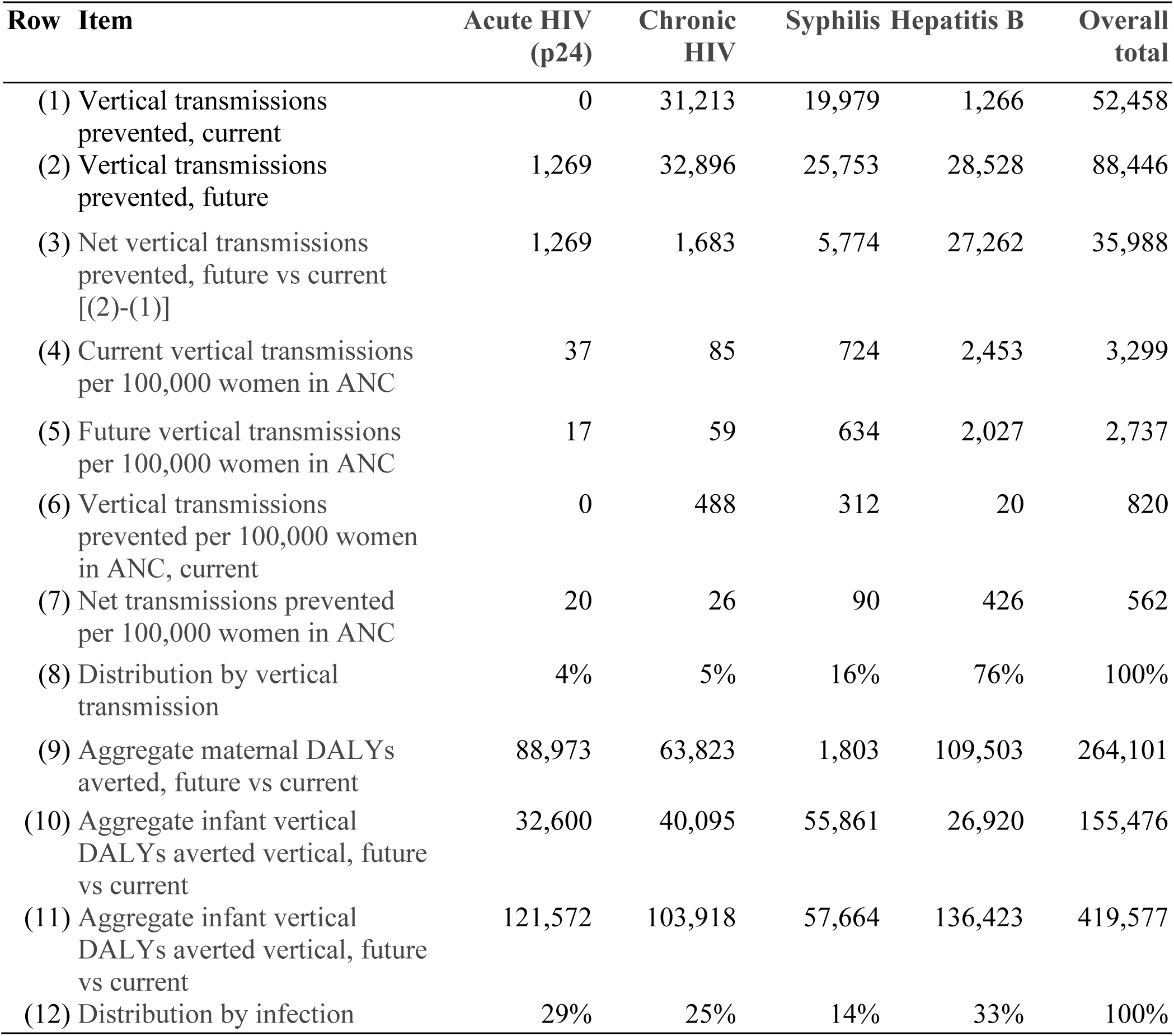
Health impacts.

The detailed results of the deterministic model quantify the contribution of p24 testing. Of the cohort of 6.400 million women in ANC, 67% (4.288 million) are tested with the ANC panel. Of these, 15,503 test positive at initial testing. The model shows these consist of 2,648 true positives and 12,855 false positives. Thus, the predicted value positive at initial testing is 17% (i.e. 2,648/15,503).

Under the best performing algorithm (provisional ART), all women who initially test positive receive provisional ART treatment for four weeks. They are asked to return to the ANC setting for confirmatory testing with Nigeria’s standard confirmatory testing (StatPak). Based on its 96.7% sensitivity and 99.2% specificity, the retesting confirms 2,558 positives; they are treated as true positives. Of the 12,855 initial false positives, StatPak correctly classifies 12,574 as negative while 231 are wrongly confirmed and remain false positives. After the confirmatory testing there are 2,790 positives (2,558 true positives and 231 false positives) so the predicted value positive rises to 92% (2,558/2,790).

### Economic benefits

The ANC testing panel entails additional testing and treatment costs (Table 3). However, in helping to prevent syphilis, the panel offsets some existing costs. Overall, the ANC panel generates net costs of $7.55 per woman. Nigeria’s public health system was expected to fund the cost of HIV testing tests done within the same panel, and the cost of treatment of HIV. Thus, $4.81. (64%) per woman would be paid publicly and only $2.74 privately. For the entire cohort of 6,400,000 women, the aggregate cost amounts to $48 million. The breakdown by infection shows that the intervention is cost-saving for syphilis (net cost of -$0.86 per woman).

**Table 3:**
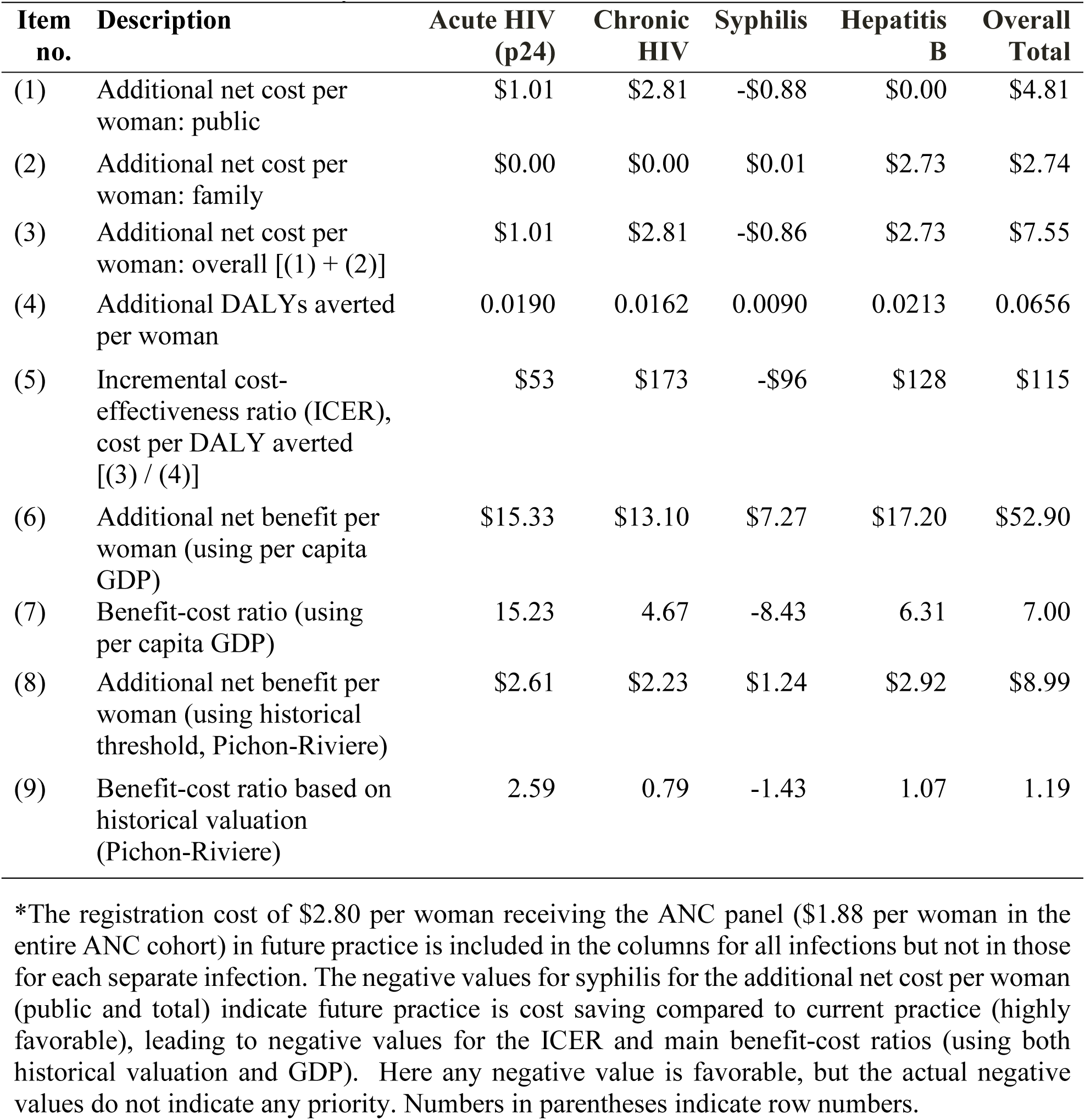
Economic results by infection*.

The overall ICER from the deterministic analysis is $115 per DALY averted. To interpret this, the most relevant threshold comes from Pichon-Rivière and colleagues[10]. They derived this threshold empirically based on the relationship of historical health spending and improvements in life expectancy and quality of life. Converted to 2025 prices, this threshold is $137 per DALY, which is far below Nigeria’s $807 GDP per capita.

To express the results as a benefit-cost ratio, we first valued each DALY averted as the per capita GDP of Nigeria. This valuation is consistent with the lower valuation of the WHO Macro-Economic Commission on Health, which considered any ICER at or below one times the country’s per capita GDP as highly cost-effective[11]. Using that threshold, we get an extremely favorable benefit-cost ratio of 7 to 1. We generated a more conservative benefit-cost ratio by valuing each DALY averted based on the country’s historical threshold of dollars per spending. The resulting benefit-cost ratio (1.19) is still quite favorable.

### Sensitivity analysis

The probabilistic sensitivity analysis shows a median ICER of $116, close the deterministic value ($115), with 95% confidence interval of $91 to $143 (see Figure 2). The probability that the ICER is below the $137 historical threshold is 91%. Thus, the results provide high confidence that the ANC panel would be cost-effective.

**Figure 2.**
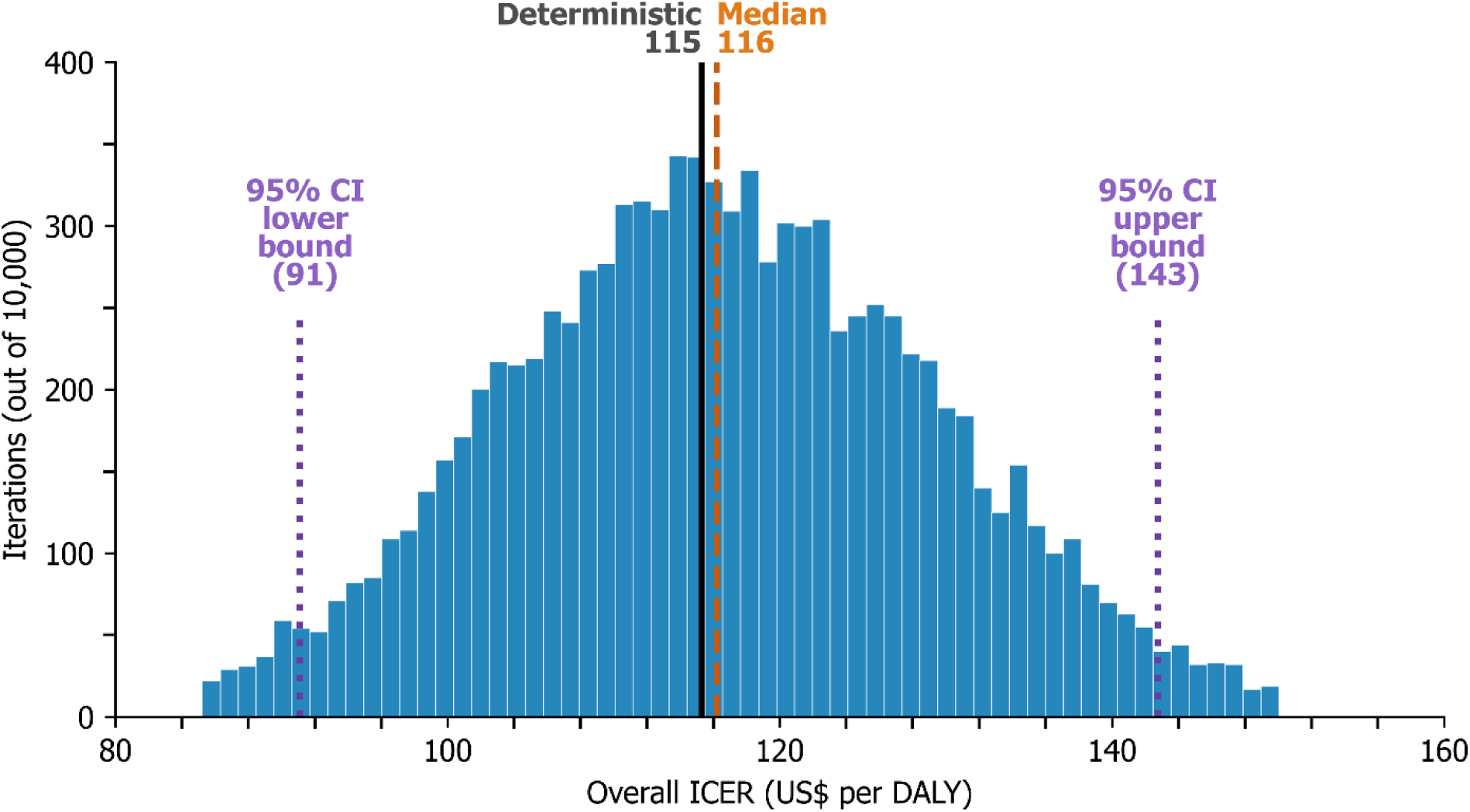
Histogram of overall ICER in probabilistic sensitivity analysis

The probabilistic sensitivity analysis gave ICER probabilities by infection of 100.0%, and the at Nigeria’s historical threshold of $137/DALY for all infections except chronic HIV (see Figure 3). Similarly, the probabilities of favorable benefit-cost ratios (at least 1.00) were also favorable for all infections except chronic HIV (see Figure 4). However, chronic HIV economic outcomes would also be favorable with 100% probability if each DALY averted were valued at Nigeria’s GDP threshold ($807).

**Figure 3.**
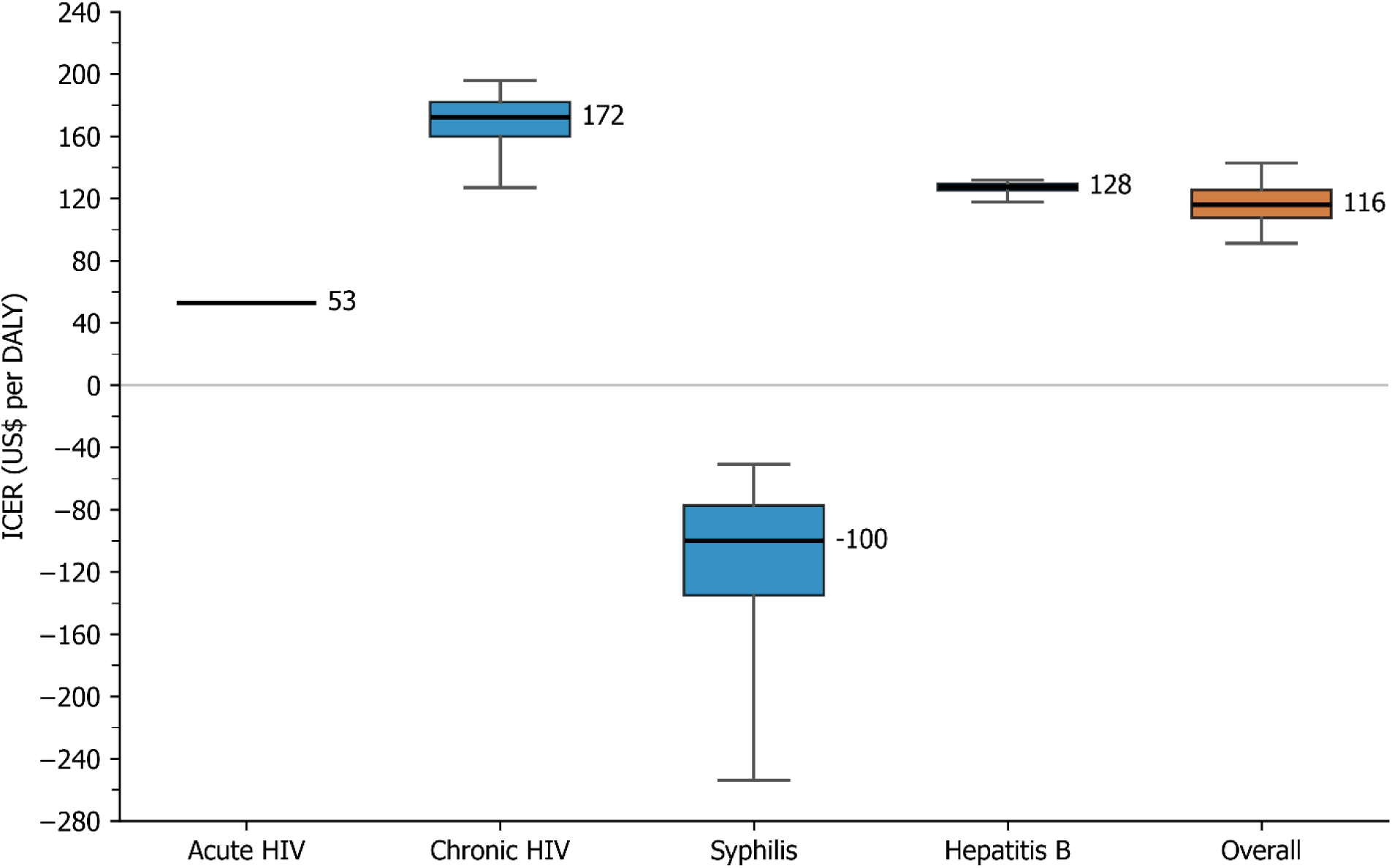
Box plot of ICER for each infection showing median, quartiles, and 95% confidence intervals from probabilistic sensitivity analysis Note: For acute HIV, there is no variability in the ICER as all costs and health impacts both depend only on the prevalence.

**Figure 4.**
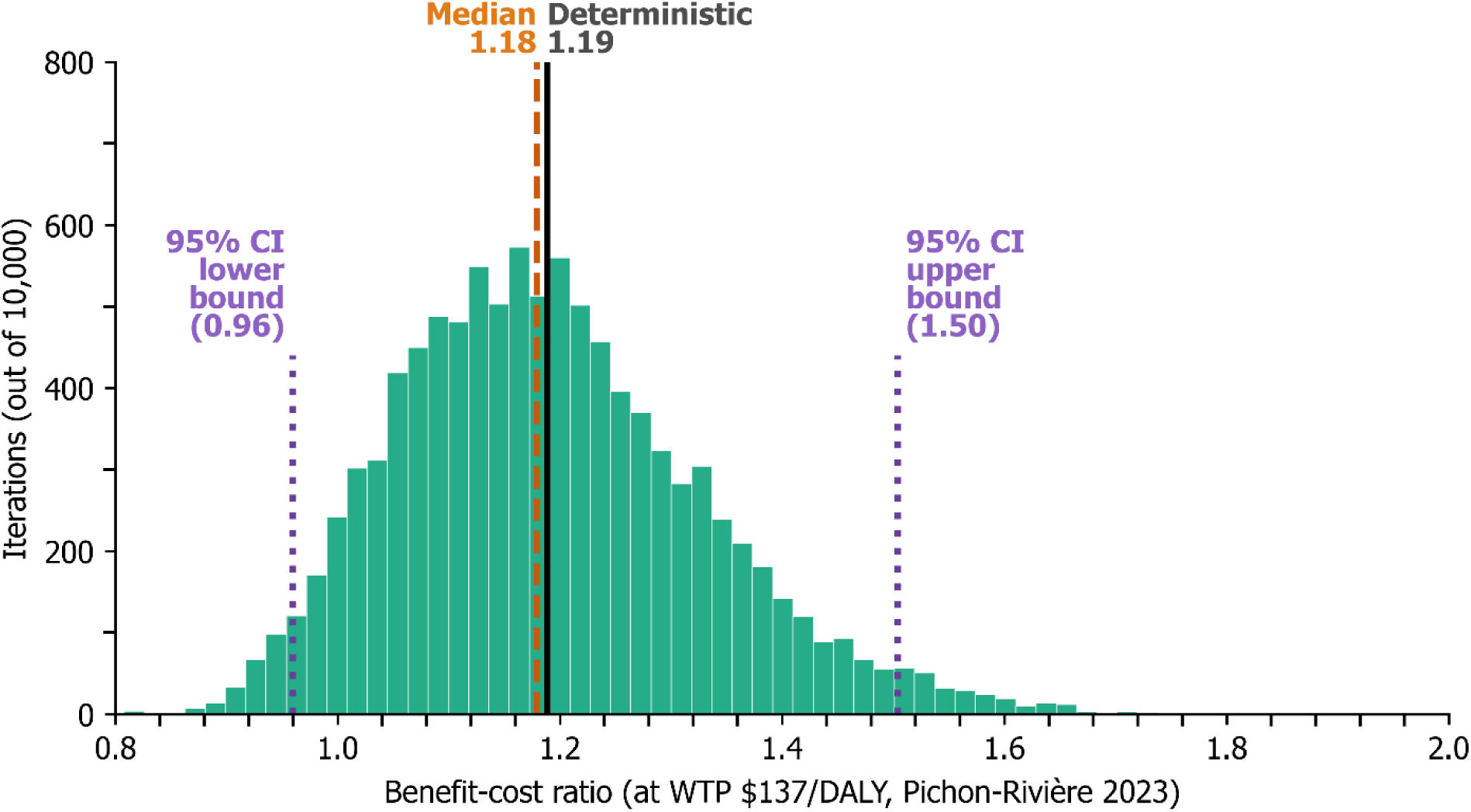
Histogram of benefit-cost ratio from probabilistic sensitivity analysis

### Alternative algorithms following positive p24

Table 4 compares the alternative algorithms. Provisional ART has the highest overall and net economic benefit per woman in ANC ($8.99 and $1.44), respectively.

**Table 4:** Summary of p24-positive algorithms (future scenario options)

| No. | Description | Provisional<br>ART | 4-week<br>return | StatPak, 4-week return | StatPak,<br>PCR | PCR<br>referral |
| --- | --- | --- | --- | --- | --- | --- |
| (1) | Program cost (\$) per woman | \$7.55 | \$7.35 | \$7.36 | \$7.41 | \$7.40 |
| (2) | Program DALYs averted per woman | 0.066 | 0.058 | 0.058 | 0.060 | 0.060 |
| (3) | Program ICER (\$/DALY) | \$115.24 | \$127.48 | \$127.57 | \$124.05 | \$123.97 |
| (4) | Economic benefit per woman (\$), historical patterns | \$8.99 | \$7.91 | \$7.91 | \$8.19 | \$8.19 |
| (5) | Benefit-cost ratio, historical [(4)/(1)] | 1.19 | 1.08 | 1.08 | 1.11 | 1.11 |
| (6) | Net economic benefit per woman (\$), historical [(4) - (1)] | \$1.44 | \$0.56 | \$0.55 | \$0.78 | \$0.79 |

Note: Numbers in parentheses in formulas refer to row numbers.

## DISCUSSION

### Rationale for selected testing algorithm

The 17% predicted value positive of the initial p24 test is the result of the high (99.7%) specificity of the p24 antigen test but the low prevalence (0.70%) of acute HIV, Provisional ART provides important protection against the adverse effects of HIV, particularly given the acute phase high transmissibility[9]. It helps protect the infant during a period of high vulnerability with minimal costs and risk of resistance to ART from excessive treatment. As ART treatment is widely available in health facilities performing ANC, feasibility is favorable and program costs per woman ($7.55) are acceptable. In PEP, the risk of acute HIV is 0.30%[12]. As noted, the predicted value positive after the initial and confirmatory tests are 17% and 92%, respectively. These are orders of magnitude higher than those used to justify PEP, indicating that the benefits of treatment substantially outweigh the risks of possibly unnecessary HIV treatment.

Under the recommended (provisional ART) algorithm, the 2,558 true positives would receive lifetime treatment, potentially saving the lives of the woman and their infants. While the 231 false positives would receive ART for several months, under standard guidelines their viral load would be measured periodically. Since they were not infected, their viral load would be undetectable at repeated visits, so that ART could then be discontinued.

Two alternatives (4-week return and Stat-Pak, 4-week return) would both miss the main benefit of prompt treatment during the period of acute vulnerability. The next two alternatives (StatPak, PCR and PCR referral), which require PCR confirmatory testing for discordant p24 results (i.e., positive on the initial screen but negative on confirmation), would require that pregnant women have regular access to PCR testing. Unfortunately, fewer than 100 facilities in Nigeria can offer public sector PCR testing, compared to the many thousands that perform HIV testing. In Nigeria, the cost of a PCR test in a private lab appears prohibitive for most women. There are a few public labs where patients could receive tests and results inexpensively, and they would generally require an on-site visit. If testing and/or feedback of results were delayed or not completed, treatment would be delayed or missed.

### Behavioral responses

Except for p24, the major benefits of the ANC panel rest in expected behavioral changes of healthcare providers and patients. While patients and providers in the Nigerian healthcare system widely engage in detecting and treating HIV, such widespread behaviors do not extend to the other infections. As Figure 1 showed, their testing rates are far lower. As the ANC testing panel performs all tests together, it operationally links all tests. Thus, the likelihood is far higher that public health system will pay for the full range of tests, the provider will recommend them, the patient will obtain them, and the woman will initiate and sustain treatment for herself and, if indicated, her infant.

### Other studies on p24 in sub-Saharan Africa and their implications

The authors have searched for other data sources on the prevalence of p24 antigen among pregnant women attending ANC. First was a 2020 study from Calabar and Cross River states in Nigeria Nigeria[13]. However, this study did not meet our quality standards as a representative sample of pregnant women in ANC (see Supplement 3).

The authors are aware of two initiatives in sub-Saharan Africa to increase understanding of acute HIV infection in ANC. The authors learned that an operations research study on p24 in Nigeria is underway. However, details on the study design or results are not available. Finally, officials from the Rwanda Biomedical Centre report that that country is staring to use p24 testing in routine antenatal care. Existing data do not distinguish reactive results between antibody or antigen positive. However, the country plans to update its procedures for recording and using the results. If future data becomes available, they will refine projections of added health benefits and added costs but are likely to have only limited impact on the ICER.

### Budget impact

Along with cost-effectiveness, budget impact impacts public health decisions. For this the proposed rollout of the ANC panel program, the bulk of the cost would be public, potentially implying a substantial budget impact for Nigeria. However, the practical budget impact would be limited. The projected net public cost per woman in ANC ($4.81) is primarily the present value of future treatment costs for HIV for the woman. Fortunately, these costs do not need to be paid up front but would be paid each year over the woman’s remaining life. This fact provides time for the federal and state health ministries to work with their respective finance ministries in arranging the necessary costs. With a woman having a discounted remaining lifetime of 20.77 years, each year’s annual treatment cost is only $0.23 (i.e., $4.81/20.77), which should be a manageable amount and critical investment in preserving the woman’s life and health.

### Extension to Sub-Saharan Africa overall

As a preliminary extension to sub-Saharan Africa, we retained our estimates of acute HIV prevalence, as the available data also came from sub-Saharan Africa. For chronic HIV we replaced the Nigerian values with overall values for sub-Saharan Africa, with no changes for other parameters. The region’s current prevalence of HIV in females of reproductive was 3.0%[14], about triple the Nigerian rate, its current testing coverage was 62.87%[15] and the current treatment coverage was 89%[16]. Future values were projected to equal future values for Nigeria. With sub-Saharan Africa’s higher prevalence, the overall DALYs averted per woman rose to 0.3407 and the net additional cost per woman rose to $31.18. The ICER changed to $91 per DALY averted and the benefit-cost ratio increased to 1.50. Although data limitations did not allow us to update all other parameters, these preliminary results suggest that the ANC panel would be comparably valuable in sub-Saharan Africa overall.

### Limitations

The main limitation is the paucity of data related to the expected positivity rate of p24 antigen in ANC. We identified only one such study from Nigeria; as noted above[13], but it did not pass the authors’ quality standards There was only one other such study from sub-Saharan Africa as a whole[8].

The second limitation was the need to rely on expert opinion for future behaviors (e.g., screening and treatment rates). While both limitations create uncertainty around aggregate costs and aggregate effectiveness, the resulting ICERs and benefit-cost ratios would not be substantially affected by alternative estimates of prevalence or testing. The reason is that aggregate costs and aggregate effectiveness are positively correlated. If the prevalence of p24, future screening rates with the ANC testing panel, or treatment rates for those infected were higher than our base estimate, more infections would be detected and treated, generating more health gains. On the other hand, more treatment entails higher cost. ICERs and benefit-cost ratios, being ratios, do not change as numerators and denominators move in similar directions.

## CONCLUSION

This analysis found that integrating the ANC panel into the health system with the assumed guidelines would be cost-effective. The largest benefits came from substantial increases in HBV testing.

The results highlight the importance of updating testing protocols to make appropriate use of the p24 capabilities to become similar to those of PEP. If those protocols are followed, recent HIV infections could be identified and treatment could be initiated promptly. However, if only non-PEP protocols designed around antibody tests were used, then ART treatment might be delayed and the health benefits substantially undermined.

While this model is calibrated with the best available data, it entails substantial uncertainty where empirical data are limited, especially around p24 prevalence and future testing and treatment rates. A demonstration project with at least 1,000 women in ANC in a country or region considering using the ANC panel would provide valuable real-world data that could refine the model estimates and inform decisions about wider adoption.

## Data Availability

Most data produced in the present work are contained in the manuscript or at the link below. All remaining items are available on reasonable request to the authors.

https://drive.google.com/drive/folders/11ht5cM82HbhGIBjz_8_mM0nO0cFFl1vk?usp=sharing

## WHAT WAS KNOWN BEFORE

- Screening of HIV, syphilis, and hepatitis in prenatal care are useful but generally done only separately and incompletely
- Coverage of chronic HIV via antibody testing is generally high.
- Coverage of syphilis and hepatitis testing and treatment in prenatal care in Nigeria and is generally low.

## WHAT THE STUDY ADDS

- Integrating syphilis and hepatitis B testing with an HIV test will substantially increase coverage
- The ANC panel test is highly cost-effective and cost-beneficial in Nigeria and likely in many other sub-Saharan countries.
- Using p24 to test for early HIV infection is a promising and cost-effective capability.

## ACKNOWLEDGMENTS

The authors thank Wisam Haddadin, Friday Iroabuchi, Michelle Lee, Aaron Mulaki, and Andrew Whitney for constructive comments on presentations and drafts, Zahibulla Nazari for assistance with literature review, and Uwayezu Ange Bertin, Charles Berabose, and Rwibasira Gallican and Kamanzi Octave Rodrigue for sharing experience with 4th-generation HIV testing in Rwanda.

## COMPETING INTERESTS

DSS and AA received funding from Abbott, Inc. for part of this study. DSS, MO and MK also received funding from Abbott for previous research. Other authors declare no conflicts.

## FUNDING

This study was funded in part by Abbott, Inc. (Abbott Park, IL, USA). The funder reviewed and commented on drafts but had no role in the study design; in the collection, analysis, and interpretation of data; in the writing or approval of the manuscript; or in the decision to submit the manuscript.

## DATA AVAILABILITY

Data are available through the sources described in this manuscript and the supplements.

## AUTHORS’ CONTRIBUTIONS

Conceptualization: Donald S. Shepard (DSS), Abdalrahman Ahmed (AA). Data curation: DSS, Pacifique Dusabeyezu (PD), Isaiah Dzimbiri (IKD), Missiani Ochwoto (MO), AA. Formal analysis: IKD. Funding acquisition: DSS. Investigation: DSS, Michael Kingsley (MK). Methodology: DSS, MK, AA. Project administration: DSS. Resources: DSS. Software: IKD. Supervision: DSS, MK. Validation: PD, IKD, MO, AA. Visualization: DSS, PD, IKD. Writing – original draft: DSS, PD, IKD. Writing – review & editing: DSS, PD, IKD, MO, MK, AA. All authors read and approved the final version of the manuscript.

## FIGURES

Google Drive Link: https://drive.google.com/drive/folders/11ht5cM82HbhGIBjz_8_mM0nO0cFFl1vk?usp=sharing

## SUPPLEMENTS

**Supplement 1:** Microsoft Excel workbook: deterministic model for Nigeria

**Supplement 2:** Python code for probabilistic sensitivity analysis (zip file with IPYNB, JSON html files)

**Supplement 3:** Data sources and interpretation for key parameters (pdf).

**Supplement 4:** Microsoft Excel workbook: deterministic model for sub-Saharan Africa

## Notes

### Competing Interest Statement

Donald S. Shepard and Abdalrahman Ahmed received funding from Abbott, Inc. for part of this study. Donald S. Shepard, Missiani Ochwoto and Michael Kingsley also received funding from Abbott for previous research. Other authors declare no conflicts.

## REFERENCES

1. WHO. Introducing a framework for implementing triple elimination of mother-to-child transmission of HIV, syphilis and hepatitis B virus. 2023. https://www.who.int/publications/i/item/9789240086784. Accessed 6 July 2026.

2. WHO, UNICEF. Country guidance for planning triple elimination of mother-to-child transmission of HIV, syphilis and hepatitis B virus programmes. 2025. https://www.who.int/publications/i/item/9789240112490. Accessed 13 June 2026.

3. Miller WC, Rosenberg NE, Rutstein SE, Powers KA. Role of acute and early HIV infection in the sexual transmission of HIV: Curr Opin HIV AIDS. 2010;5(4):277–282.

4. Routy J-P, Cao W, Mehraj V. Overcoming the challenge of diagnosis of early HIV infection: a stepping stone to optimal patient management. Expert Rev Anti Infect Ther. 2015;13(10):1189–1193.

5. IAS 2025, the 13th IAS Conference on HIV Science. 2025. https://www.iasociety.org/conferences/ias2025. Accessed 13 June 2026.

6. Umoke M, Sage P, Bjoernsen T, Umoke PCI, Ezeugworie C, Ejiofor D, et al. Co-infection and risk factors associated with STIs among pregnant women in rural health facilities in Nigeria: a retrospective study. Inq J Health Care Organ Provis Financ. 2021;58:0046958021992912.

7. Shepard DS, Halasa-Rappel YA, Rowlands KR, Kulchyckyj M, Basaza RK, Otieno ED, et al. Economic analysis of a new four-panel rapid screening test in antenatal care in Kenya, Rwanda, and Uganda. BMC Health Serv Res. 2023;23(1):815.

8. Ochwoto M, Matiang’i M, Machuki Onchieku N, Ndoria S, Matoke L, Otinga M, et al. The feasibility and impact of deploying a four-tests panel at antenatal care in primary health care facilities of a developing country, Kenya. Front Public Health. 2024;12:1399612.

9. Federal Ministry of Health, National Agency for the Control of AIDS. Nigeria HIV/AIDS Indicator and Impact Survey (NAIIS) 2018: National Summary Report. 2019. Abuja, Nigeria. Federal Ministry of Health https://naca.gov.ng/wp-content/uploads/2021/02/NAIIS-PA-NATIONAL-FACTSHEET-FINAL.pdf. Accessed 13 June 2026.

10. Pichon-Riviere A, Drummond M, Palacios A, Garcia-Marti S, Augustovski F. Determining the efficiency path to universal health coverage: cost-effectiveness thresholds for 174 countries based on growth in life expectancy and health expenditures. Lancet Glob Health. 2023;11(6):e833–e842.

11. Bertram M, Lauer J, De Joncheere K, Edejer T, Hutubessy R, Kieny M-P, et al. Cost– effectiveness thresholds: pros and cons. Bull World Health Organ. 2016;94(12):925–930.

12. Vento S. HIV transmission in sub-Saharan Africa: excessive focus on heterosexual vaginal intercourse. Front Public Health. 2025;13:1667141.

13. Ndeh FJ, Ojong EW, Akpan UO, Ekeagba II. Prevalence of human immunodeficiency Virus p24 antigenemias and mother -to- child transmission rate among HIV 1 & 2 antibody- sero-negative apparently healthy pregnant women in Calabar, Nigeria. 2020;10(12). 10.5281/zenodo.4415357.

14. World Bank. World Bank Indicators. 2024. https://data.worldbank.org/indicator/SH.DYN.AIDS.ZS?locations=ZG. Accessed 6 July 2026.

15. Raru TB, Merga BT, Deressa A, Birhanu A, Ayana GM, Negash B, et al. Coverage and determinants of HIV testing and counseling services among mothers attending antenatal care in sub-Saharan African countries: a multilevel analysis. BMC Public Health. 2024;24(1):910.

16. World Bank. Antiretroviral therapy coverage for PMTCT (% of pregnant women living with HIV). 2024. https://data.worldbank.org/indicator/SH.HIV.PMTC.ZS. Accessed 6 July 2026.

